# Trust in healthcare providers, information sources, and concerns for new maternal vaccines among pregnant and lactating women in Kenya

**DOI:** 10.1101/2025.03.24.25324583

**Authors:** Jessica L. Schue, Berhaun Fesshaye, Emily Miller, Prachi Singh, Molly Sauer, Rosemary Njogu, Rose Jalang’o, Joyce Nyiro, Ruth A. Karron, Rupali J. Limaye

## Abstract

New maternal vaccines have the potential to reduce morbidity and mortality for infants from common illnesses that pose the greatest risk in the earliest phase of their life. Respiratory syncytial virus (RSV) is a leading cause of acute lower respiratory infections among infants under six months of age. With the recent approval of a maternal vaccine for RSV, this study aimed to understand decision-making factors among pregnant and lactating women for receiving a newly licensed vaccine during pregnancy. Pregnant and lactating women from two counties in Kenya, Nakuru and Mombasa, were recruited to complete a cross-sectional survey in August-September 2022. The survey explored topics of trust in various types of sources for information about new maternal vaccines, the importance of a healthcare provider’s recommendation of a new maternal vaccine, and concerns about new maternal vaccines. We surveyed 400 pregnant and lactating women. In both counties, information about the new vaccine was most trusted when coming from healthcare providers, and least trusted when coming from social media. Women’s intention to receive a new maternal vaccine was heavily influenced by a positive recommendation from a healthcare provider. The greatest concerns about a new vaccine were side effects and the vaccine’s ingredients. The information and recommendation from a healthcare provider are important influences on decision-making for new maternal vaccines. As a new maternal immunization for RSV becomes more available, healthcare providers should be engaged early to reduce vaccine hesitancy amongst providers and equip providers with appropriate information tailored to pregnant women about the RSV maternal vaccine.

## Introduction

Maternal immunization, or immunization during pregnancy, allows the pregnant person to pass antibodies through the placenta to the fetus, thereby providing passive immunity that protects infants at birth and in the first weeks and months of life (1). Vaccination during pregnancy can also serve to protect the pregnant person from experiencing severe disease or death themselves, and maternal infections are themselves a contributor to preterm birth which can subsequently contribute to neonatal mortality (2,3).

The first vaccine to be administered during pregnancy was tetanus toxoid to prevent neonatal tetanus (4). Since then, tetanus-diphtheria (Td) or tetanus-diphtheria-pertussis (Tdap) and SARS-CoV2/COVID-19 vaccines have been added to the list of maternal immunizations that are now commonly implemented worldwide (4). In high- and middle-income countries, influenza and most recently, respiratory syncytial virus (RSV) vaccines are recommended for pregnant women, whereas these vaccines are not standard in low- and middle-income countries (5,6). Looking ahead, several maternal Group B Streptococcus (GBS) vaccine candidates are currently in development to help overcome the limitations of existing prevention strategies and better protect newborns from infection (7), and maternal immunization against malaria and tuberculosis is also being considered.

Respiratory syncytial virus (RSV) is the leading cause of acute lower respiratory infections in children under five, and in 2019, was estimated to cause more than 33 million infections, 3.6 million hospital admissions, and more than 100,000 deaths (8). Low- and middle-income countries (LMICs) bear a disproportionate burden of the disease, accounting for 95% of infections and 97% of deaths (8).

A bivalent RSVpreF maternal vaccine has been approved for use and recommended to pregnant women in more than 40 high- and middle-income countries as of the end of 2024, with the World Health Organization recommending that all countries introduce either the maternal vaccine or a long-acting monoclonal antibody to protect infants against RSV disease (9,10). Unlike maternal RSV vaccines, long-acting RSV monoclonal antibodies are administered to infants to provide protection for several months, but are costly with extremely limited supply, leaving them out of reach in all but higher-income settings (11,12). Given the limited availability of RSV monoclonal antibodies in LMICs, the maternal RSV vaccine is particularly important. The RSV maternal vaccine is recommended for use during the third trimester of pregnancy, with recommendations for the exact gestational age windows varying by country (9,13).

Previous research in Kenya has shown a substantial burden of RSV-related illness and death among children under 5 years of age, with highest levels of morbidity and mortality occurring in the first 6 months of life (14). While non-medically attended RSV disease accounts for most of the country’s burden, RSV hospitalization is also a significant cause of economic burden for households in Kenya (15,16). This makes preventative care like vaccination in pregnancy especially important as it can not only decrease RSV-related morbidity and mortality, but also decrease economic burden in Kenya.

Given that a maternal RSV vaccine may be available in Kenya in the next few years, this study sought to understand which information sources pregnant women trusted and their intentions toward future maternal vaccines given in pregnancy.

## Methods

Participants were recruited from 20 health facilities in Nakuru County (rural) and Mombasa County (urban). A range of health facility types was included, from level 2 (community clinics) through level 5 (specialized referral hospitals).

Data collectors completed a standard training, and the survey instrument was pretested. Recruitment procedures differed by facility; generally, participants were approached consecutively upon arrival at antenatal clinics, maternity wards, or maternal and child health units. If a participant met the inclusion criteria (18+, currently breastfeeding or in the second or third trimester of pregnancy, and able to consent), oral consent was obtained, with a place on the consent form for the participant to mark an X to indicate their consent. Surveys were administered in either English or Swahili using tablets. Data collection occurred from 26 July 2022 through 30 sept 2022. This study received ethical approval from the Kenya Medical Research Institute Scientific and Ethics Review Unit (protocol 4211) in Kenya and Johns Hopkins Bloomberg School of Public Health Institutional Review Board (study IRB00014893) in the US.

The survey instrument included sociodemographic questions, trust in information sources, the importance of a healthcare provider’s recommendation, and concerns about new maternal vaccines. The survey started with questions about RSV risk perception and specific questions about an RSV maternal vaccine, followed by questions about new maternal vaccines broadly.

### Trust in information sources

We asked women to indicate their level of trust in a variety of sources, asking participants: “I trust the information that I have received from xx about vaccines during pregnancy.” We asked about the following sources: health care provider, social media platforms, scientists and doctors at universities and academic institutions, and media (TV, radio), and women were given a 4-point Likert scale for answer choices (strongly agree, agree, disagree, strongly disagree).

### Healthcare provider recommendation

We asked women to indicate their likelihood of receiving a new vaccine based upon their healthcare provider’s recommendation. We asked: “If a new vaccine was approved for pregnant women, and your health care provider did not recommend it, how likely would you get the new vaccine?” and “If a new vaccine was approved for pregnant women, and your health care provider did recommend it, how likely would you get the new vaccine?”. Women were given a 4-point Likert scale for answer choices (very likely, likely, unlikely, very unlikely).

### New vaccine concerns

We asked question about future vaccine concerns and asked participants to rank their answers from 1-6, with 1 being most concerning and 6 being least concerning: “When a new vaccine is approved for use and is recommended for me/family, I am typically concerned with the following: ingredients in the vaccine; side effects; availability of the vaccine at my health facility; costs to get the vaccine; what others are saying about the vaccine; healthcare providers recommendation to get the vaccine; a family member, such as a partner or mother-in-law’s input about me getting the vaccine.”

## Results

We surveyed 400 participants, evenly split between Nakuru and Mombasa counties. The majority of participants were between the ages of 18-29, had at least one child, and nearly half of participants had at least a secondary education (Table 1).

**Table 1:** Sociodemographic characteristics of study sample.

|  | *Nakuru*<br>(*N=200*) | *Mombasa*<br>(*N=200*) | *p-value* |
| --- | --- | --- | --- |
| Age, n(%) |  |  |  |
| *18-29* | 132 (66.0) | 137 (68.5) | 0.594 |
| *30-44* | 68 (34.0) | 63 (31.5) |  |
| Lactating, n(%) |  |  |  |
| *Pregnant* | 51 (25.5) | 50 (25.0) | 0.908 |
| *Lactating* | 149 (74.5) | 150 (75.0) |  |
| Number of children under 18, n(%) |  |  |  |
| *None* | 19 (9.5) | 20 (10.0) | 0.556 |
| *One* | 63 (31.5) | 69 (34.5) |  |
| *Two* | 66 (33.0) | 58 (29.0) |  |
| *Three* | 28 (14.0) | 36 (18.0) |  |
| *Four or more* | 24 (12.0) | 17 (8.5) |  |
| Education level, n(%) |  |  |  |
| *Less than primary school* | 15 (7.5) | 30 (15.0) |  |
| *Primary school completed* | 68 (34.0) | 62 (31.0) |  |
| *Secondary/ High school completed* | 66 (33.0) | 55 (27.5) |  |
| *College/University completed* | 51 (25.5) | 53 (26.5) | 0.097 |

In both Nakuru and Mombasa, 98% of respondents agreed or strongly agreed that they would trust information about new maternal vaccines in pregnancy provided by healthcare providers. A majority of respondents in both counties also trusted information from scientists and/or doctors and media, including TV and radio. Trust in information from social media was low in both counties, as 78% and 80% for Mombasa and Nakuru respectively disagreed that they could trust social media (Figure 1).

**Fig 1:**
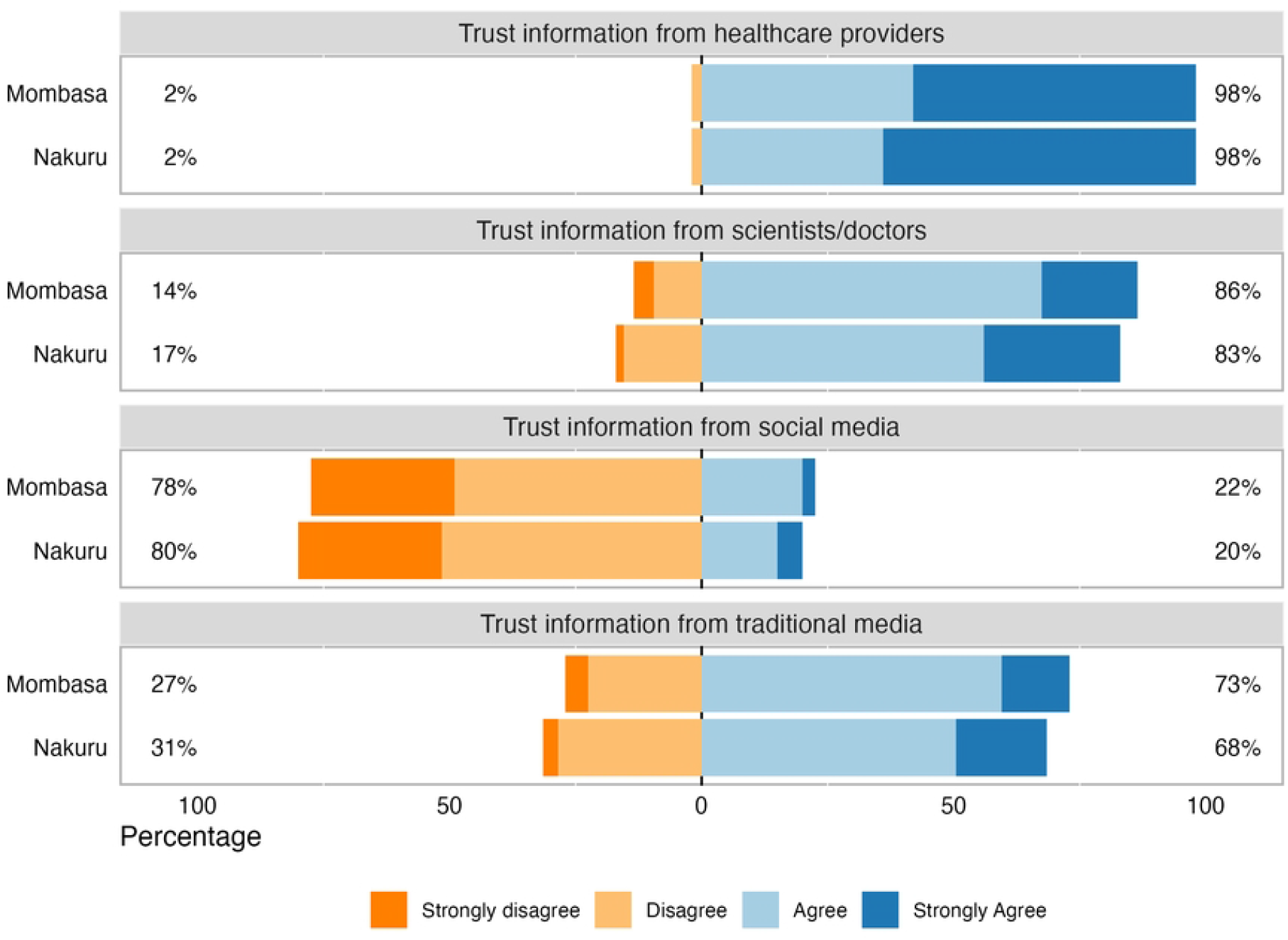
Level of trust in information about new maternal vaccines by source No respondents chose Strongly Disagree in response to trusting information from healthcare providers.

We then asked participants about healthcare provider recommendations. If their healthcare provider recommended a new maternal vaccine, 63% of women in Nakuru and 70% of women in Mombasa indicated that they were very likely to get the new maternal vaccine. If their healthcare provider did not recommend a new maternal vaccine, 5% of women in each county indicated that they were very likely to get the new maternal vaccine (Figure 2).

**Fig 2:**
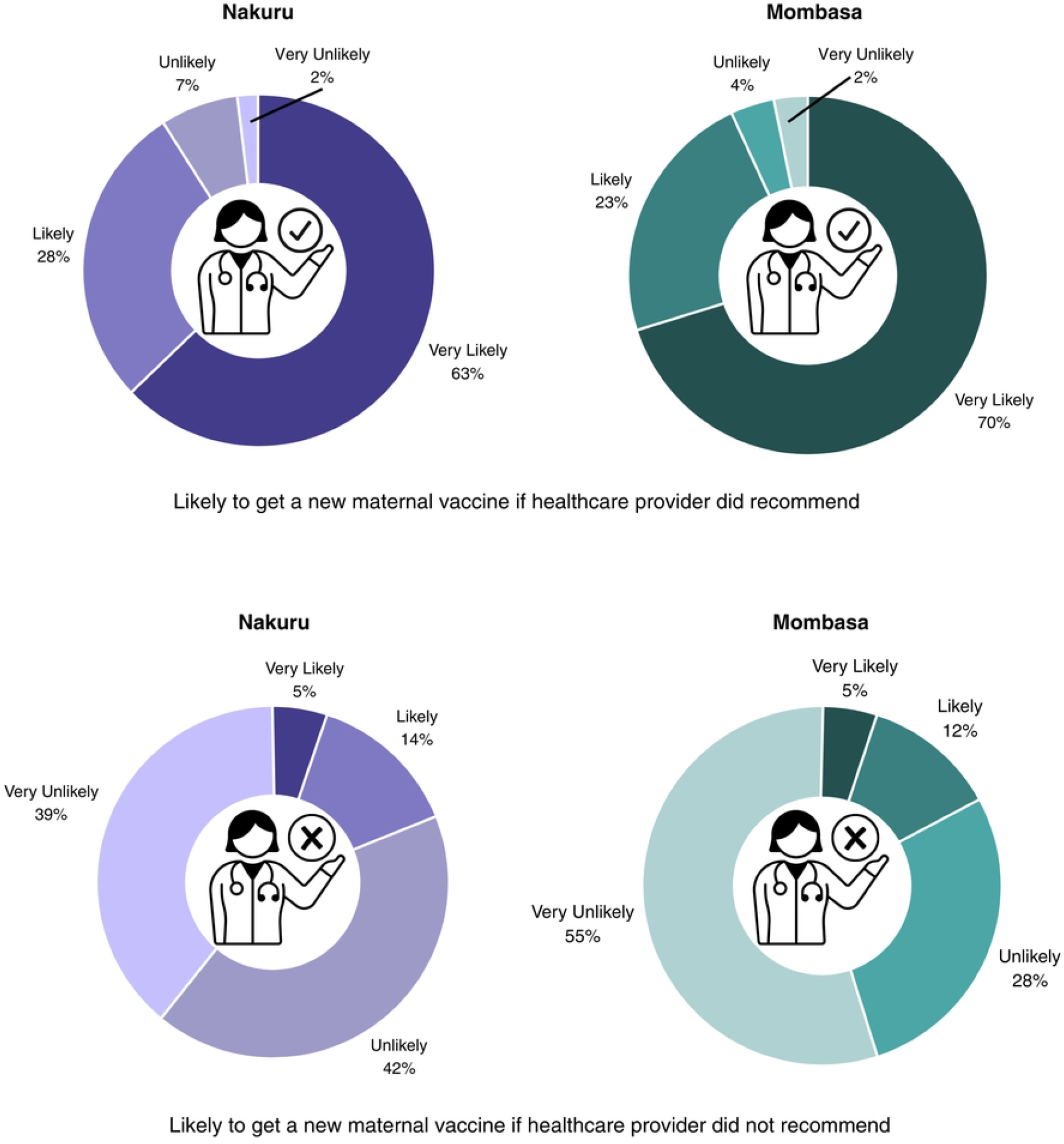
Influence of healthcare provider recommendations on maternal vaccine acceptance

Related to concerns about new maternal vaccines, we examined how participants ranked concerns by county of residence and displayed these in a heatmap. The concern that was most frequently ranked as the most important across both counties was vaccine side effects, with the second concern being the vaccine’s ingredients. In both counties, opinions of family members and others were the lowest ranked concerns. (Figure 3).

**Fig 3:**
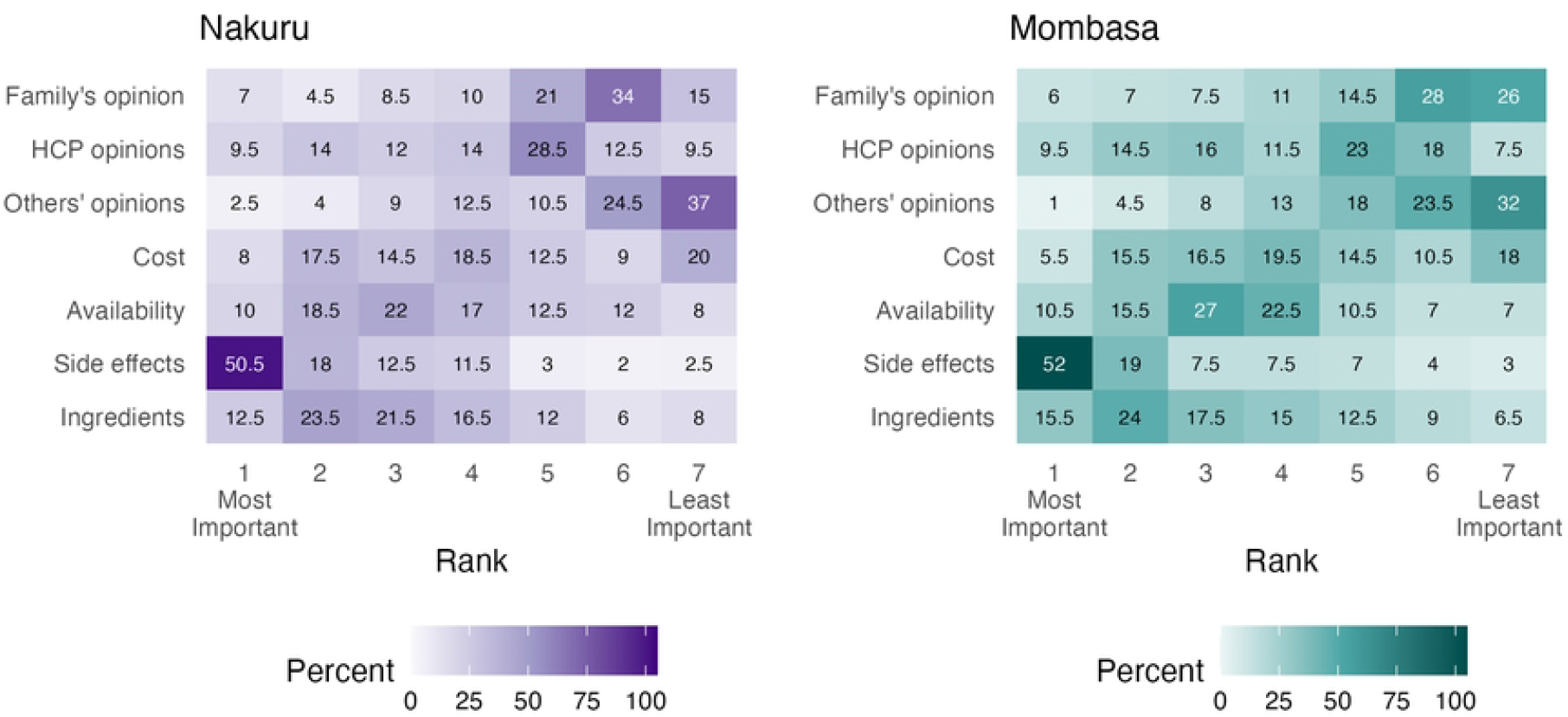
Concerns about new maternal vaccines ranked Values and shades of tiles represent the percent of respondents that chose a particular concern at each rank level. HCP: Healthcare provider.

## Discussion

Healthcare providers were highly trusted sources for information about new maternal vaccines among pregnant and lactating women in both Nakuru and Mombasa counties in Kenya. A healthcare provider’s recommendation to receive or not receive a new maternal vaccine holds considerable influence on women as they decide whether to be vaccinated in pregnancy. The main concerns about a new maternal vaccine are the vaccine’s side effects and ingredients. Participants were least concerned about their family’s and others’ opinions about them getting a new maternal vaccine. Responses were similar between the two counties, suggesting negligible difference in opinions between urban and rural populations. While these specific responses were about new maternal vaccines broadly, the questions were prompted by describing maternal RSV vaccines.

While research on maternal RSV vaccines in low- and middle-income countries is lacking, there have been a few studies focused on knowledge and acceptance of RSV vaccines in pregnancy. A study among Australian women focused on RSV and Group B Streptococcus (GBS) vaccines in pregnancy found that safety data was the most important factor for acceptance (17). These findings align with ours as side effects, an important measure for safety, was the biggest concern among women in both counties. A sample of women in Jordan also found favorable opinions about RSV vaccination during pregnancy, given the vaccine is safe (18). The concern of vaccine ingredients, which emerged as the second most important in both counties, was not as prevalent in other studies.

Previous research, including from Kenya, has shown that healthcare providers are important and highly trusted sources for health information, including vaccination in pregnancy (19–21). Aligned with our findings, a recommendation for vaccination from a healthcare provider is central to vaccine acceptance, and the absence of a recommendation can even decrease uptake (19,20). One study also found that pregnant women even preferred to speak with healthcare providers directly about vaccination, instead of receiving printed communication materials (22). Similar to our results, two previous studies in Kenya also found that traditional media sources such as TV and radio, were utilized and trusted more than social media sources (23,24). However, despite the potential presence of misinformation on social media, some studies have found social media and Internet platforms as important sources of health information for mothers (25,26). This points to the importance of ensuring accurate information is present on social media to increase trust and present correct information to those that are already relying on these platforms.

The growing evidence base on maternal vaccination acceptance highlights the importance of targeted and tailored communication strategies, particularly those that equip healthcare providers to deliver clear and consistent information to their patients throughout pregnancy. Safety concerns, like side effects or concerns about vaccine ingredients, are commonly identified in similar studies. Strengthening surveillance systems to monitor and evaluate adverse events among pregnant women and newborns is one strategy to improve the information available to providers and program officials to guide recommendations and healthcare provider recommendations (27). Distinguishing serious AEFI from local vaccine reactions, and understanding how common they are, may also help build trust and mitigate concerns about side effects and ingredients. For example, relatively minor side effects (i.e., pain at the injection site, swelling, etc.) from tetanus vaccination in pregnancy have not deterred vaccine acceptance (28). Discussing these experiences may help providers assuage patients’ concerns and foster greater acceptance of future maternal vaccines.

RSV vaccine is recommended in the third trimester, a relatively narrow window compared to other vaccines in pregnancy (i.e., tetanus toxoid) which may be given at any time in pregnancy. A study in Kenya found that two in three pregnant women had their first antenatal care (ANC) visit prior to this window (29). This may lay the groundwork for healthcare providers to prepare pregnant women for newer maternal vaccines by providing information and addressing concerns during their first ANC visits, before they are eligible to receive these vaccines. Our study found that media is a trusted information source among pregnant and lactating women. Targeted media and social media campaigns in advance of RSV vaccine availability should include messages emphasizing the opportunity for mothers to protect their newborns by being vaccinated themselves, and including information about vaccine safety as well as efficacy (30).

This study had several limitations. First, participants were exclusively recruited from health facilities, so the opinions of women who do not seek care through the formal health system are missed. In addition, social desirability bias may have influenced participants to respond with more favorable views of vaccines or healthcare providers than they actually believed. Despite these limitations, the study also had notable strengths. Participants were recruited from both urban and rural settings in different parts of the country. This is also one of the first studies to address information needs for future maternal vaccines, which is crucial for demand generation and community sensitization.

Pregnant and lactating women rely on trusted sources to inform their decision-making related to maternal vaccines. As our study revealed that a strong recommendation from a healthcare provider would likely be the most important influence, it will be imperative to equip healthcare providers with salient and digestible information to be able to strongly recommend a future maternal RSV vaccine. Healthcare providers can also be vaccine hesitant, and efforts to provide relevant information to them well before a maternal RSV vaccine is available is therefore important to realize the potential benefits of a maternal RSV vaccine at the population level.

## Data Availability

Data will be made available upon reasonable request.

